# Validity of resting heart rate derived from contact-based smartphone photoplethysmography (PPG) compared with electrocardiography (ECG): Protocol for a Systematic Review and Meta-Analysis

**DOI:** 10.1101/2025.06.13.25329618

**Authors:** James D Mather, Nicholas F Sculthorpe, Nilihan E.M. Sanal-Hayes, Ethan Berry, Jacqueline L. Mair, Lawrence D. Hayes

**Author notes:** **Correspondence:** Nilihan E.M Sanal-Hayes. Co-author emails: James Mather, Nicholas Sculthorpe, Ethan Berry, Jacqueline Mair, Lawrence Hayes.

## Abstract

**Objective:** To synthesise quantitative evidence on the validity of photoplethysmography (PPG) derived from mobile devices (i.e. smartphones) for the assessment of heart rate (HR) compared to the gold standard electrocardiogram (ECG).

**Introduction:** Mobile health (mHealth), the use of mobile devices for promotion or measurement of health is on the rise. Smartphone cameras can perform photoplethysmography (PPG) for the assessment of heart rate (HR) and other cardiac cycle characteristics. However, rigorous validity is necessary before smartphone measurement of PPG can be utilized for healthcare provision. There is a pervasive belief that HR-PPG is analogous to HR-ECG, and herein we will provide an updated systematic review and meta-analysis to support or challenge this supposition.

**Inclusion criteria:** This review will include studies that investigate the correlation coefficient between of resting heart rate (RHR) acquisition from PPG utilizing contact-based smartphone devices versus ECG as the gold standard, Studies will be excluded if they (a) do not use PPG utilizing contact-based smartphone devices (b) compare PPG to another collection method other than ECG, or (c) are review articles or case studies.

**Methods:** A comprehensive literature search will be conducted in CINAHL Ultimate, MEDLINE, ScienceDirect, and Scopus using a search strategy developed in collaboration with the research team. All retrieved citations will be imported into Rayyan for screening and data management. A minimum of two independent reviewers will conduct the title and abstract screening, followed by two independent reviewers who will perform full-text screening and data extraction. All stages will be guided by predefined inclusion and exclusion criteria, which will be pilot tested to ensure consistency and reliability. Any discrepancies will be resolved through discussion with a third reviewer or during a research team meeting. Intra-rater reliability will be quantified at the title and abstract stage, and the full-text review stage using Cohen’s Kappa. To ensure clarity and consistency in the presentation of study characteristics and findings, both narrative synthesis and tabular formats will be employed.

**Review registration:** https://doi.org/10.17605/OSF.IO/83V7A

## Introduction

Photoplethysmography (PPG) has been widely employed over several decades for the diagnosis, monitoring, and screening of various diseases and disorders, offering clinically relevant physiological insights [1–4]. The term “photoplethysmography” derives from its functional components: “photo” (light), “plethysmo” (volume), and “graphy” (recording) [5]. Initially introduced by Hertzman in 1937 to detect blood volume changes [4,6], PPG operates by measuring either transmitted (transmissive PPG) or reflected (reflective PPG) light as it interacts with biological tissues [7]. This technique relies on the optical properties of tissue, including absorption, scattering, and transmission [8]. Transmissive PPG detects light that has passed through relatively thin tissue regions, such as the fingers, toes, or earlobes. In contrast, reflective PPG captures light that is scattered back from the skin, which results in a reduction in detected light intensity [9]. While transmissive PPG generally provides more stable signal quality[10], reflective PPG offers greater versatility in terms of measurement site, enabling its application in anatomical regions such as the forehead, wrist, carotid artery, and esophagus— locations where transmissive PPG is less feasible [11–14].

Photoplethysmography (PPG) operates based on the Beer–Lambert Law, which describes the attenuation of light intensity as a function of the extinction coefficient, concentration of the absorbing medium, and the optical path length through which light travels [15]. Leveraging this principle, a range of PPG devices are utilized in clinical settings to measure physiological parameters such as pulse rate, a key vital sign [7]. Clinically, PPG is commonly employed to monitor cardiac-induced fluctuations in blood volume within microvascular beds at peripheral anatomical sites including the finger, forehead, earlobe, and toe [16,17].

Since the introduction of the first iPhone in 2007, smartphones have become ubiquitous worldwide and are increasingly recognized as practical tools for data collection, addressing several limitations inherent in traditional methods [18]. Conventional health monitoring typically involves periodic, scheduled clinical visits, which may fail to capture dynamic physiological changes that occur longitudinally or during routine daily activities [3,19,20]. In this context, smartphones equipped with integrated cameras offer a cost-effective alternative for PPG acquisition, eliminating the need for additional external devices such as wearables [21].

Consequently, smartphone-based PPG has the potential to extend access to underserved populations, particularly those facing demographic, geographic, or socioeconomic barriers to healthcare access and delivery [22–24]. The adoption of mobile health (mHealth) technologies has accelerated in recent years, particularly following the COVID-19 pandemic, which underscored the utility of remote and prospective health and symptom monitoring [21,25–27]. As such, the increasing prevalence of smartphone-enabled telemedicine is likely to persist and may play a significant role in advancing global health equity, aligning with targets outlined in the United Nations Sustainable Development Goals (UN SDGs), specifically SDG 3: Good Health and Well-being [28,29].

Smartphone PPG can estimate RHR through the measurement of distal pulse rate (PR) at rest, and whilst completing other activities such as exercise or cognitive tasks [7]. However, to convert the PPG signal to an estimate of HR, a mathematical algorithm is required which may influence validity and reliability. These algorithms are rarely available in validation studies, limiting transparency. This is problematic given the proliferation of mHealth, and therefore the necessity that technologies are reliable and valid compared to gold standard measurements prior to universal adoption [30]. To examine validity of smartphone-derived HR (HR-PPG), De Ridder *et al.* [31] meta-analyzed studies published until 7^th^ December 2016 examining HR-PPG compared to several methods including ECG, pulse oximetry, and radial pulse. Results revealed good agreement between HR-PPG and validated method-derived RHR. Authors concluded that RHR obtained from HR-PPG could be an alternative to traditional methods but only in an adult population, and only in the right context. De Ridder and colleagues [31] highlighted several limitations to the included studies. Firstly, there was high statistical heterogeneity between studies, ostensibly due to participant characteristics, measurement conditions, and the smartphone devices utilized. Secondly, the latest iOS device reviewed was the iPhone 5 (released 2012) and the latest android was the Samsung Galaxy S4 (released 2013). Emerging evidence suggests advancements in technology, such as the availability of various camera positions (i.e., front-facing vs rear-facing) and the advent of multiple lenses could result in improvements in PPG acquisition [21]. Our recent scoping review [3] revealed ten studies that compared HR-PPG to ECG, and we reported extensive methodological study characteristics and guidelines for researchers and practitioners aiming to optimize HR-PPG. However, we did not quantify agreement between HR-PPG and ECG. Therefore, we thought it pragmatic to provide an update on the validity of HR-PPG compared to ECG, quantifying the literature using meta-analytical techniques.

Review questions:

This review poses primary and secondary research questions:

Primary research question: What is the agreement between HR-PPG and ECG?

Secondary research questions:

1. Are there differences in the agreement of HR-PPG based on device parameters (e.g., iOS or Android operating systems)?
2. What is the risk of bias and methodological quality of studies investigating agreement between HR-PPG and ECG?

We approach this review with the hypothesis that HR-PPG may offer therapeutic a valid alternative to ECG for measurement of resting hear rate. We also expect that, the evidence base will be limited but emerging, given the recency of smartphone devices for telemedicine and the time required to conduct and publish controlled trials, and most studies will be small-scale or pilot RCTs, possibly with methodological variability in study design, and data collection parameters. Given the novelty and evolving use of HR-PPG, we anticipate some heterogeneity in how populations are defined across studies, which may influence both inclusion decisions and the strength of conclusions.

### Eligibility Criteria

#### Participants

This review will investigate studies with measurement of HR-PPG via front or rear facing camera of a smartphone by contact-based PPG. Only studies which report a correlation coefficient between HR-PPG and the gold standard measurement (ECG) will be included.

We will exclude articles if the index measurement was conducted with a device connected to a smartphone, such as a mobile sensor, medical device or wearable device; the paper did not include validity assessment of HR-PPG and HR-ECG as an outcome measurement; the study used a clinical population (we assumed healthy population unless stated otherwise); the paper was not an original article (i.e., utilized a database from a secondary source); the paper was a review; there was no abstract or full text available.

#### Types of Sources

This review will consider articles that present at least pilot data and include quantitative data. Review articles, case studies or qualitative studies will therefore not be considered.

## Methods

This meta-analysis will be conducted in accordance with the PRISMA (Preferred Reporting Items for Systematic Reviews and Meta-Analyses) guidelines. The protocol has been registered with Open Science Framework: https://doi.org/10.17605/OSF.IO/83V7A. Any deviation from the protocol will be clearly logged with dates, reasons, and impacts noted.

### Search strategy

Our search strategy was designed to achieve a balance between comprehensive coverage of the relevant literature and practical constraints, while maintaining a high level of scientific rigour. We selected four databases—CINAHL Ultimate, MEDLINE, ScienceDirect, and Scopus—based on their relevance to the interdisciplinary nature of our topic, which spans health sciences, data processing, and behavioural research. CINAHL Ultimate was chosen for its strong coverage of allied health literature. MEDLINE (accessed via EBSCOhost) provides authoritative biomedical literature, including cardiac-focussed studies. ScienceDirect was included due to its extensive full-text access to journals in signal processing, physics, and related disciplines published by Elsevier. Scopus was selected for its comprehensive indexing of interdisciplinary research and citation tracking capabilities, allowing us to identify additional relevant studies. We will search grey literature alongside peer-reviewed sources.

This includes preprints (e.g., MedRXiv, arXiv), dissertations (via ProQuest), conference proceedings (e.g., Web of Science), and institutional repositories like DSpace. Moreover, we will supplement database searches by reviewing reference lists, identifying citing articles via tools such as CrossRef and Google Scholar, and using CoCites to find articles with similar citation patterns. We will validate our search by testing it against known relevant studies to ensure key articles are captured. If the initial database search yields a high proportion of irrelevant records, the search strategy (including keywords, subject headings, and Boolean logic) will be iteratively refined to improve precision and relevance. This process will continue until the strategy demonstrates adequate sensitivity and specificity. In cases where additional information is required to assess study eligibility or to extract necessary data, we will contact the corresponding authors using a standardized email template. If no response is received, reminder emails will be sent at two and four weeks following the initial contact. All attempts to contact authors, as well as any responses received, will be documented in detail. To enhance transparency, we will report the total number of authors contacted, the number who responded, and the number who provided the requested data. This information will be summarized in the main manuscript, with a supplementary table detailing the nature of the information requested and received. Although this will not be a living systematic review, we will re-run the database searches prior to submission if more than six months have elapsed since the initial search.

Query strings:

(((((“validity”) AND (“mobile”)) AND (“photoplethysmography”)) OR (“PPG”)) AND (“heart rate”)) AND (electrocardiogram OR ECG OR EKG) NOT (“wearable”) AND (2007:2025[pdat]).

The dates have been selected as 2008 was the year the first Apple iPhone was released, so to ensure we have captured all articles, we started the search in 2007 until the end of 2025.

This review focuses on correlation coefficients between HR-PPG and HR-ECG. We will also gather data on devices used, study participants, location of study, commercial availability of application, sampling rate, camera position and resolution, flash (torch) settings, channel used for computations, ECG device utilized, electrode placement, ECG processing information, instructions given to participants, dietary control, participant posture, region of interest, breathing pattern, environmental conditions, stabilization period, duration of measurement, and number of attempts or trials.

### Study/Source of Evidence Selection

After the search, all citations will be uploaded and have their duplicates removed in Rayyan [32]. The screening process will consist of two rounds: (1) title and abstract screening, and (2) full-text screening. In both rounds, two independent reviewers will screen each record using pre-specified inclusion and exclusion criteria developed *a priori*. Blinding of reviewers during screening will be implemented to the extent possible using Rayyan, which allows for independent decisions without visibility into the other reviewer’s judgments. This helps reduce bias and increases objectivity in the initial phases of study selection. Conflicts between reviewers will be flagged automatically by Rayyan and subsequently resolved through discussion with two other reviewers. If consensus cannot be reached, a third reviewer (independent chair) will adjudicate. To assure consistency and transparency, all reviewers will undergo a calibration exercise using a small sample of studies before formal screening begins. At both the titles and abstracts stage and the full text stage, inter-rater reliability will be calculated and expressed via Cohen’s Kappa statistic, scores range from –1 to 1 with scores closer to 1 indicating stronger agreement. This exercise will help refine the application of the inclusion/exclusion criteria and ensure a shared understanding of borderline cases. We will share the full list of sources from the database searches and screening decisions by individual screeners. Bibliographic data (titles, abstracts, metadata) will be exported in RIS and CSV formats, while screening decisions will be provided in a separate CSV/XLSX file. All files will be uploaded to an open-access repository, like OSF, upon manuscript submission or acceptance.

### Data Extraction

During the training and calibration phase, a small subset of studies (approximately 5–10%) will be independently extracted by all reviewers using a draft version of the data extraction form. Discrepancies will be discussed and used to refine the data extraction protocol and ensure consistency across reviewers.

In the primary data extraction phase, two reviewers will independently extract predefined data items using a standardized and piloted extraction form. Extracted variables will include, but are not limited to, correlation coefficients, photoplethysmography (PPG) parameters, electrocardiogram (ECG) parameters, participant demographics, and device-specific characteristics. During the risk of bias and methodological quality assessment phase, two reviewers will independently evaluate study-level risk of bias using validated tools, such as the Cochrane Risk of Bias tool. Any disagreements will be resolved through consensus discussion or adjudicated by a third reviewer if necessary. In the reconciliation and verification phase, extracted data will be compared, and discrepancies will be resolved through discussion or third-party adjudication. Finalized data will then be entered into the meta-analysis dataset. Where appropriate, artificial intelligence (AI) or computer-assisted tools may be used to support the identification of relevant text passages or extraction of bibliographic metadata. However, all outputs from such tools will be verified by human reviewers. All stages of the data extraction and risk of bias assessment process will be conducted or supervised by human reviewers to ensure methodological rigor and data accuracy.

Data extraction will follow PRISMA guidelines and Cochrane Handbook procedures, using the standardized form provided in the OSF project. Only data related to HR-PPG and HR-ECG correlation coefficients, as defined by the study authors, will be extracted, with missing data marked as “NR” and flagged for follow-up.

Primary study information, including bibliographic details (author(s), year, title, journal,), and setting (e.g., clinical, academic), will be recorded. Data extracted from each study will include sample size, participant sex, country of study, age, skin pigmentation, if participants were considered healthy, smartphone model, name of application utilized, whether the application was commercially available, index measurement sampling rate, camera position and resolution, flash (torch) settings, channel used for computations, ECG device utilized, electrode placement, and ECG processing information.

Statistical data (e.g., effect sizes) will be extracted. Risk of bias will be assessed using an appropriate tool if found. Ambiguous data will be flagged for discussion, and assumptions or clarifications will be noted in comments, with no estimation of missing data unless explicitly instructed. All numerical entries will be double-checked prior to submission, and a second reviewer will verify the data, resolving discrepancies through discussion or adjudication by a third reviewer. Completed forms will be saved and uploaded to the OSF folder following the naming convention: StudyID_ExtractorInitials_Extraction.xlsx.

Each round of data extraction will be conducted by two independent extractors working in parallel, covering study characteristics, participant data, and risk of bias assessments. Extractors will receive standardised training and work independently, with results compared and discrepancies reconciled through discussion. A third reviewer will adjudicate if disagreements persist. Inter-rater reliability will be assessed during training using Cohen’s kappa. These results will inform adjustments to the extraction form and training. In each data extraction round, two independent extractors will compare their entries side by side, with differences highlighted automatically. They will then discuss discrepancies, referring to source material and instructions, to reach consensus. If they cannot agree, a third senior reviewer will adjudicate, reviewing the relevant documents and providing a final decision, recorded in the reconciliation log. All discrepancies and resolutions will be documented. Systematic discrepancies will be analysed to update the extraction protocol if needed. Once reconciled, the data will be finalised for analysis and included in the meta-analytic dataset.

### Data Analysis and Presentation

The analysis will be carried out using the Fisher r-to-z transformed correlation coefficient as the outcome measure. A random-effects model will be fitted to the data, with analyses conducted in R Studio using the *metafor* package. Weltz et al. [33] proposed that when conducting a meta-analysis involving Fisher r-to-z transformed correlation coefficients, especially in the presence of study heterogeneity, employing a random-effects model with appropriate variance estimation techniques yields more reliable and generalizable results compared to fixed-effects models. Therefore, random effects models will be utilized. Studentized residuals and Cook’s distances will be used to examine whether studies may be outliers and/or influential in the context of the model [34]. Studies with a studentized residual larger than the 100 × (1 – 0.05/(2 × *k*))th percentile of a standard normal distribution will be considered potential outliers (i.e., using a Bonferroni correction with two-sided α = 0.05 for *k* studies included in the meta-analysis). Studies with a Cook’s distance larger than the median plus six times the interquartile range of the Cook’s distances will be considered to be influential. The rank correlation test [35] and the regression test [36], using the standard error of the observed outcomes as predictor, will be used to check for funnel plot asymmetry. Heterogeneity will be assessed via *I²*, *Cochran’s Q*, and *tau^2^* statistics. Subgroup analyses will explore variations in HR-PPG validity based on operating system, participant characteristics (e.g., age, sex), and study features if possible. Moderator analyses will assess how PPG parameters and participant traits influence outcomes, and the impact of risk of bias will be tested through sensitivity analyses. Additional sensitivity checks will examine the effects of excluding high-risk studies, handling missing data, and methodological quality. Where available, follow-up data will be used to assess treatment durability, and adverse effects will be summarised qualitatively or quantitatively. If data allow, meta-regression will be used to examine relationships between continuous variables and effect sizes. Results will be presented using forest and funnel plots, and a GRADE profile will summarise the quality of evidence for each outcome. Conclusions will be based effect sizes with overall effect size r>0.9 (approximately z>1.5) being considered valid. For reference, a value of r=0.70 is generally considered very large, and this is approximately equal to z=0.8. High heterogeneity (*I²* > 50%) will prompt cautious interpretation and exploration through subgroup or moderator analyses. The GRADE framework will guide the confidence in conclusions, depending on evidence quality.

## Discussion

This meta-analysis protocol aims to provide an updated quantification of the validity of HR-PPG compared to HR-ECG in healthy subjects at rest. With the rapid development in technology and an improved understanding of this research area, this will be an important finding, indicating whether HR-PPG can be used with confidence. There is a pervasive belief that HR-PPG is analogous to HR-ECG, and herein we will provide an updated systematic review and meta-analysis to support or challenge this supposition.

The decision to conduct a meta-analysis arose from the need to identify agreement of HR-PPG with HR-ECG, including modifying parameters such as frame rate, camera location, skin pigmentation, operating system. Moreover, if there is an absence of knowledge around modifying variables, this systematic review will provide opportunities for future research. By providing context to current findings, this research can guide future studies to concerning PPG and its application.

### Project Timeline

The estimated project timeline includes screening of titles and abstracts is from September 2025 to November 2025. Screening of full texts is estimated to be from November to December 2025. Results are expected by February 2026. During this period, we will update our database searches to identify any newly published studies that meet our inclusion criteria.

## Acknowledgements

None.

## Data availability

This is a protocol paper, so no data are currently available. Once the study is complete, the dataset supporting the findings will be included in the main manuscript and Appendix.

## Authors contributions

All authors contributed equally to the conceptualisation, preliminary search, search strategy, writing – original draft, review and editing

